# Short telomeres, but not accelerated epigenetic or proteomic age, increase fibrotic lung disease risk: Mendelian randomisation with pleiotropy dissection and independent replication

**DOI:** 10.64898/2026.10.14.26364960

**Authors:** Sten Hanke

## Abstract

**Background:** Short telomeres are an established cause of idiopathic pulmonary fibrosis (IPF), but the genetic evidence rests on seven instruments, six of which lie at telomere-maintenance loci, and so cannot separate an effect of telomere length from an effect of the telomerase pathway itself — a distinction that determines whether telomere length is a tractable target or only a marker of the pathway. Whether other measures of biological ageing share the effect, and whether accelerated ageing causes immune-mediated disease more generally, has not been tested systematically.

**Methods:** Two-sample Mendelian randomisation of six ageing measures — GrimAge, PhenoAge, Hannum-age and intrinsic epigenetic age acceleration, proteomic age acceleration, and leukocyte telomere length — against six immune-mediated diseases in FinnGen R12: giant cell arteritis (GCA), polymyalgia rheumatica, primary Sjögren’s disease, interstitial lung disease (ILD), Crohn’s disease and ulcerative colitis. All 36 pairs were analysed in both directions using inverse-variance weighted (IVW), MR-Egger, weighted-median and weighted-mode estimators, with Cochran’s *Q*, *I*², leave-one-out and single-variant diagnostics, a pre-specified analysis excluding HLA-region instruments, and iterative *Q*-based outlier removal. Instruments were annotated against telomere-maintenance and pulmonary-fibrosis loci and that class excluded. Minimum detectable effects were computed for every pair before estimates were examined. Any pair surviving correction was re-estimated in two independent IPF case-control datasets. A two-step analysis tested mediation of ILD risk by seven gut microbial taxa acting through telomere length.

**Results:** One of 36 forward pairs survived correction. Longer telomeres reduced ILD risk: outlier-robust IVW odds ratio (OR) 0.59 per standard deviation (SD), 95% CI 0.49–0.71, *p* = 2.7 × 10⁻⁸, on 77 instruments, with no residual heterogeneity (*I*² = 6%) and concordance across estimators. Four pleiotropic instruments — in *TERT*, *TERC*, *APOE* and *ATM* — inflated the unfiltered estimate to OR 0.46 (0.37–0.57) and accounted for all of its heterogeneity. The effect was reproduced in 5,159 clinician-diagnosed European IPF cases (OR 0.18, 0.12–0.25, *p* = 5.0 × 10⁻²³) and in 1,210 East Asian cases (OR 0.54, 0.35–0.83, *p* = 0.005). Excluding every telomere-maintenance and pulmonary-fibrosis locus left OR 0.26 (0.17–0.40), *p* = 1.6 × 10⁻⁹, on 72 instruments, establishing that the effect does not depend on *TERT* or *TERC*. The effect was three times larger for clinically defined IPF than for the broad registry ILD endpoint (*p* = 1.8 × 10⁻⁹), consistent with dilution of a fibrosis-specific mechanism by case mix. No methylation clock and no proteomic age measure affected any disease; smallest detectable ORs ranged from 1.29 to 5.24 per SD, so these nulls exclude only large effects. The reverse direction could not be tested at informative precision. One apparent clock–autoimmunity association was produced entirely by a single HLA-region instrument. Mediation was null at its first step.

**Conclusions:** Telomere attrition causally increases fibrotic lung disease risk, with an effect that concentrates in clinically defined IPF and that survives removal of every telomere-maintenance locus — shown here for the first time — and is therefore attributable to telomere length rather than to the telomerase genes that dominate conventional instrument sets. Four methylation clocks and proteomic age acceleration show no such effect, so these measures cannot be treated as interchangeable proxies for biological ageing.

## Introduction

Giant cell arteritis is the commonest primary systemic vasculitis in adults over 50 in European populations, essentially does not occur below that age, and has a mean onset near 75 years [13,14]. Polymyalgia rheumatica overlaps it clinically and shares its age distribution. This steep age dependence, with the inflammatory pathophysiology of both, makes accelerated biological ageing an attractive causal explanation, and similar reasoning is applied to primary Sjögren’s disease, connective-tissue-disease-associated interstitial lung disease, and the inflammatory bowel diseases.

Biological age can be proxied genetically. DNA-methylation clocks estimate biological age from blood methylation: Hannum-age [7] and Horvath’s pan-tissue clock [16] were trained to reproduce chronological age, whereas PhenoAge [17] and GrimAge [8,18] were trained against mortality and morbidity, making GrimAge the measure most strongly associated with age-related disease. Leukocyte telomere length indexes replicative ageing inversely [2]. Proteomic age acceleration derives from plasma protein panels [3]. Genome-wide association studies exist for all of these [1,2,3], so each can serve as an exposure in Mendelian randomisation, where genetic variants act as instrumental variables. Because genotype is fixed at conception, MR estimates are not confounded by lifestyle or socioeconomic position, and reverse causation can be probed by exchanging exposure and outcome.

Two gaps motivate this study. First, the age-dependent vasculitides have never been tested with an ageing exposure. Second, the epigenetic-clock MR literature is large and growing, yet clock instruments include variants in the HLA region — the dominant susceptibility locus for most immune-mediated disease — placing the independence and exclusion-restriction assumptions directly at risk for exactly these outcomes. Whether published clock–immune associations survive removal of those instruments is not routinely reported.

We therefore tested all 36 combinations of six ageing measures and six immune-mediated diseases in both directions, with HLA and outlier sensitivity analyses throughout and explicit detection limits for every pair, and asked finally whether gut microbial composition acts on disease through biological ageing.

## Methods

Reporting follows the STROBE-MR recommendations [21] and the community expectations for MR analyses set out in [25]; the completed 20-item checklist is supplied.

### Study design

Two-sample MR using published summary statistics; no individual-level data. The three core assumptions are relevance, independence, and exclusion restriction. Relevance was assessed by the per-instrument *F*-statistic; independence was addressed by the pre-specified HLA sensitivity analysis; exclusion restriction was probed by comparing estimators with differing pleiotropy assumptions, by *Q*-based outlier detection, and — for the one positive finding — by annotating instruments against loci with a known direct route to the outcome.

### Exposure data

Full summary statistics were used throughout. GrimAge, PhenoAge, Hannum-age and intrinsic epigenetic age acceleration (IEAA, the Horvath-clock residual adjusted for blood cell composition) came from the European-ancestry arm of a 28-cohort meta-analysis (*N* ≈ 34,450–34,470) [1]. Proteomic age acceleration came from a UK Biobank plasma-proteomic analysis (*N* = 44,435) [3]. Leukocyte telomere length came from UK Biobank (*N* = 472,174) [2]. Sources and accessions are in Table 1.

**Table 1.** Data sources.

| Role | Trait | Source | Accession / endpoint | Sample |
| --- | --- | --- | --- | --- |
| Exposure | GrimAge acceleration | McCartney et al. [1], European arm | GCST90014288 | 34,467 |
| Exposure | PhenoAge acceleration | as above | GCST90014292 | 34,463 |
| Exposure | Hannum-age acceleration | as above | GCST90014289 | 34,449 |
| Exposure | Intrinsic EAA (Horvath) | as above | GCST90014290 | 34,461 |
| Exposure | Proteomic age acceleration | Mörseburg et al. [3] | — | 44,435 |
| Exposure | Leukocyte telomere length | Codd et al. [2], UK Biobank | — | 472,174 |
| Outcome | Giant cell arteritis | FinnGen R12 [4] | M13_GIANTCELL | 1,387 / 484,260 |
| Outcome | Polymyalgia rheumatica | FinnGen R12 [4] | M13_POLYMYALGIA | 4,992 / 484,260 |
| Outcome | Primary Sjögren's disease | FinnGen R12 [4] | M13_SJOGREN | 3,309 / 484,260 |
| Outcome | Interstitial lung disease | FinnGen R12 [4] | ILD_ENDPOINTS | 5,765 / 494,583 |
| Outcome | Crohn's disease | FinnGen R12 [4] | K11_CD_STRICT2 | 2,489 / 497,622 |
| Outcome | Ulcerative colitis | FinnGen R12 [4] | K11_UC_STRICT2 | 7,220 / 492,160 |
| Supplementary outcome | Temporal arteritis | FinnGen R12 [4] | M13_TEMPARTERIT | 1,225 / 484,260 |
| Mediator exposure | 7 gut microbial taxa | Kurilshikov et al. [5] | GCST90016940-GCST90017065 | 6,363-17,859 |
| Replication outcome | IPF, European, clinician-diagnosed | Chin et al. [23], 7 case-control collections | GCST90984755 | 5,159 / 27,459 |
| Replication outcome | IPF, East Asian | Zhou et al. [24], GBMI East Asian arm | GCST90399719 | 1,210 / 254,409 |

### Outcome data and sample overlap

Outcomes were FinnGen R12 endpoints [4]: `M13_GIANTCELL` (GCA; 1,387 cases / 484,260 controls), `M13_POLYMYALGIA` (4,992 / 484,260), `M13_SJOGREN` (3,309 / 484,260), `ILD_ENDPOINTS` (5,765 / 494,583), `K11_CD_STRICT2` (Crohn’s disease; 2,489 / 497,622) and `K11_UC_STRICT2` (ulcerative colitis; 7,220 / 492,160). M13_TEMPARTERIT (temporal arteritis; 1,225 / 484,260) was analysed as a supplementary GCA endpoint, interpreted at nominal α and excluded from correction because it shares most of its cases with `M13_GIANTCELL`.

Exposure and outcome samples are largely but not entirely distinct. The proteomic-age [3] and telomere-length [2] exposures derive from UK Biobank, which does not contribute to FinnGen. The clock meta-analysis [1] includes one Finnish contributing cohort (the Finnish Twin Cohort), so a small overlap with FinnGen cannot be excluded; it is confined to a single cohort within a 34,000-participant meta-analysis set against a 500,000-participant outcome biobank. We did not attempt a quantitative overlap correction, as the participant-level cohort membership required is not public. Residual bias from overlap interacting with instrument strength acts toward the null in a two-sample design with strong instruments, so it is unlikely to generate false positives.

Outcome estimates were read from full summary-statistic files rather than from browser query interfaces, which thin non-significant variants and would bias instrument retrieval.

### Instrument selection

Variants with *p* < 5 × 10⁻⁸ were retained, restricted to biallelic single-nucleotide variants with minor allele frequency ≥ 0.01 and, for proteomic age, imputation INFO ≥ 0.8. Independent instruments were selected by retaining the lowest-*p* variant per 10,000 kb window, matching the window conventionally used for linkage-disequilibrium clumping. No LD reference panel was available, so residual correlation within a window could not be evaluated; because the procedure discards genuinely independent variants inside each window it is conservative with respect to independence and costs power rather than validity. This is why the telomere-length analysis uses fewer instruments than the number of independent loci reported by the source GWAS [2].

### Harmonisation

Exposure and outcome effect alleles were aligned on the reference/alternative pair of the outcome dataset. Palindromic variants were oriented by allele frequency and dropped when the exposure frequency fell between 0.42 and 0.58. Of 154 independent instruments, 141 were carried into analysis. All 13 losses were in the telomere-length set (94 → 81): five strand-ambiguous palindromes and eight absent from the outcome dataset. No instrument was lost for the other five exposures. Harmonisation was verified rather than assumed: across retained instruments the exposure effect-allele frequency correlated with the aligned outcome frequency at *r* = 0.98, with no variant differing by more than 0.2.

### Causal estimation and scale

The primary estimator was multiplicative random-effects IVW. Three estimators with weaker assumptions were run alongside: MR-Egger [9], whose intercept tests directional pleiotropy; the weighted median [10], valid when at least half the instrument weight is valid (1,000 parametric bootstrap replicates); and the weighted mode-based estimator [19] (φ = 1, 1,000 replicates, median-absolute-deviation standard error). Heterogeneity was quantified by Cochran’s *Q*, *I*² and, for NOME assessment, *I*²_GX. Leave-one-out IVW estimates and single-variant Wald ratios were computed for every pair.

Effects are reported **per standard deviation of the exposure**, the only scale on which the six measures are comparable and the scale the power analysis uses; for the four methylation clocks the per-year value is also given, that being the published scale. The exposure standard deviation was recovered from the data by comparing the analytic standard error under a standardised exposure with the observed fixed-effect standard error. This was internally consistent to within 2% across the seven outcomes for every exposure (Table 2), and the recovered values for the clocks (3.7–5.2 years) agree with those published, supporting the procedure.

**Table 2.** Instruments and detection limits by exposure. Mean and minimum *F* are computed over the instruments used in analysis.

| Exposure | Independent | Used | Mean $F$ | Min $F$ | Variance explained | Recovered SD | Detectable OR per SD |
| --- | --- | --- | --- | --- | --- | --- | --- |
| GrimAge acceleration | 4 | 4 | 37.1 | 31.1 | 0.43% | 3.96 y | 2.08-5.24 |
| PhenoAge acceleration | 11 | 11 | 51.3 | 31.8 | 1.64% | 5.17 y | 1.45-2.34 |
| Hannum-age acceleration | 9 | 9 | 48.3 | 30.8 | 1.26% | 3.75 y | 1.53-2.63 |
| Intrinsic EAA (Horvath) | 22 | 22 | 56.0 | 31.1 | 3.57% | 4.20 y | 1.29-1.78 |
| Proteomic age acceleration | 14 | 14 | 53.6 | 30.5 | 1.69% | 2.78 | 1.45-2.31 |
| Leukocyte telomere length | 94 | 81 | 161.5 | 30.7 | 2.77% | 0.99 | 1.33-1.92 |

Detectable OR is the range across the six primary outcomes at 80% power, Bonferroni-corrected for 36 tests.

### Sensitivity analyses

Three sensitivity analyses were applied.

*HLA exclusion.* Every IVW estimate was repeated after excluding instruments in chr6:28.4–33.5 Mb (GRCh38), the extended HLA region.

### Outlier-robust estimation

Iterative Cochran’s *Q* outlier removal was applied, following the per-instrument *Q*-contribution approach of radial Mendelian randomisation [22]: at each iteration the per-instrument contribution to *Q* was referred to a χ²₁ distribution with a Bonferroni threshold within the instrument set, flagged instruments were removed, and the estimate recomputed until stable. A floor of three retained instruments was imposed, because unconstrained removal reduced one four-instrument set to a single variant and produced a spurious significant result. This procedure follows the outlier logic of MR-PRESSO but is not that test, and is reported as an iterative *Q*-based procedure rather than as MR-PRESSO; no formal distortion test is provided.

#### Locus annotation for the positive finding

All telomere-length instruments were annotated against telomere-maintenance and pulmonary-fibrosis genes (*TERT*, *TERC*, *RTEL1*, *OBFC1/STN1*, *NAF1*, *PARN*, *TINF2*, *ATM*, and the chr19 *ZNF208* telomere-length region) within 500 kb, and the analysis repeated with that class excluded.

Directionality was assessed by comparing the variance each instrument explains in the exposure with that in the outcome, using Wald *t*-statistics with the effective sample size for binary outcomes. This is an approximation to, not an implementation of, the published Steiger test [27] and is reported as such.

### Multiple testing and power

Four testing families were specified: the 36 forward pairs (Benjamini–Hochberg), the 36 reverse pairs (Benjamini–Hochberg), the supplementary temporal-arteritis endpoint (nominal α), and the mediation arm (Bonferroni across seven taxa). The Bonferroni-equivalent threshold within each 36-pair family is 0.05/36 = 1.39 × 10⁻³.

For every pair the minimum detectable effect at 80% power was computed analytically from the variance in the exposure explained by its instruments, the case and control counts and the significance threshold [20]. For the reverse direction, where the outcome is continuous and the exposure is a disease liability, detection limits were derived from the achieved standard error, which makes no assumption about the scale of liability. Detection limits are reported alongside every estimate so that each null can be read as informative or uninformative.

### Mediation

A two-step design tested gut taxon → ageing measure → disease using seven candidate taxa from a 24-cohort gut-microbiome GWAS [5] (genera *Akkermansia*, *Bacteroides*, *Bifidobacterium*, *Faecalibacterium*, *Ruminococcus gnavus* group; families *Lachnospiraceae*, *Ruminococcaceae*). Taxon instruments used the locus-wide threshold *p* < 1 × 10⁻⁵ conventional for microbiome MR, pruned as above. Because these summary statistics carry no allele-frequency column, palindromic variants were dropped outright rather than oriented. Mediated effects were obtained by the product of coefficients with a delta-method standard error. The minimum detectable mediated effect was computed before any estimate was examined.

### Replication

Any pair surviving correction was taken forward to replication in outcome datasets sharing no participants with either the discovery outcome sample (FinnGen) or the exposure sample (UK Biobank). Candidate datasets were enumerated from the GWAS Catalog for idiopathic pulmonary fibrosis and interstitial lung disease, and screened on cohort composition reported in the source publications. Datasets built on UK Biobank or FinnGen were excluded as circular, which removed several otherwise well-powered deposits.

Two replication samples met the criterion. The first is a meta-analysis of seven clinician-diagnosed IPF case-control collections of European ancestry — Colorado, US, UK, IPF-JES, Genentech, UUS and CleanUP-UCD — comprising 5,159 cases and 27,459 controls, with cases diagnosed by a respiratory clinician according to international consensus criteria and no biobank electronic-health-record case source [23]. The second is the East Asian arm of a multi-biobank IPF meta-analysis, 1,210 cases and 254,409 controls [24], which is independent of both discovery samples by ancestry as well as by cohort and therefore also tests transportability of the effect beyond European ancestry.

Replication used the same instruments, the same estimators and the same sensitivity cascade as discovery, with two modifications. For the East Asian sample, palindromic instruments were dropped outright rather than oriented by allele frequency, because allele frequencies differ genuinely between ancestries and so cannot adjudicate strand. Positions were matched on rsID, both replication datasets being distributed on GRCh38 while the exposure data are on GRCh37. Effect estimates for the two case definitions — the broad registry ILD endpoint used in discovery and clinically defined IPF — were compared with a two-sided *z*-test on the difference of log odds ratios, treating the samples as independent.

Independence from the discovery samples is not the same as independence from the prior literature. The European replication meta-analysis [23] subsumes the case-control collection that served as the replication cohort of an earlier telomere–IPF Mendelian randomisation [26], so our European IPF estimate is not independent of that study’s. It differs from it in using 80 instruments rather than seven, a larger case set, and a locus-class sensitivity analysis that the seven-variant design could not support. We report it as a better-instrumented re-estimation, not as an independent confirmation. The East Asian sample [24] is independent of all prior work in this area.

Because instruments are selected in a European-ancestry exposure GWAS, the cross-ancestry replication additionally assumes that variant–exposure effects are similar in East Asian ancestry. That assumption is not testable here and is treated as a limitation, not as established.

### Software

Analyses were performed in Python 3.13 using NumPy 2.3 and SciPy 1.16. Estimators were implemented for this study following the published definitions cited above rather than taken from an existing package; weighted-median and weighted-mode standard errors used 1000 bootstrap replicates and the mode bandwidth parameter was φ = 1. The implementation was validated against the analytic standard-error relationship described under *Causal estimation and scale*, which it reproduced to within 2%. All instrument tables, harmonised datasets, per-estimator results, diagnostics and analysis scripts are provided (Data and code availability).

## Results

### Instruments

The six exposures yielded 154 independent instruments, 141 of which entered analysis (Table 2): 4 for GrimAge acceleration, 11 for PhenoAge, 9 for Hannum-age, 22 for IEAA, 14 for proteomic age and 81 for telomere length. Mean per-instrument *F*-statistics over the instruments used were 37–162, with a minimum of 30.5, so no instrument was weak. Instruments explained 0.43% (GrimAge) to 3.57% (IEAA) of variance in their exposure. Recovered exposure standard deviations were 3.7–5.2 years for the methylation clocks, 2.78 units for proteomic age, and 0.98–0.99 for telomere length, indicating that the telomere source file is approximately standardised.

### Leukocyte telomere length and interstitial lung disease

Of 36 primary forward pairs, one survived correction (Figure 1a). The outlier-robust estimate is **OR 0.59 per SD (95% CI 0.49–0.71), *p* = 2.7 × 10⁻⁸** on 77 instruments, with no residual heterogeneity (*Q p* = 0.32, *I*² = 6.3%) and concordance across estimators (weighted median 0.59, weighted mode 0.79).

**Figure 1.**
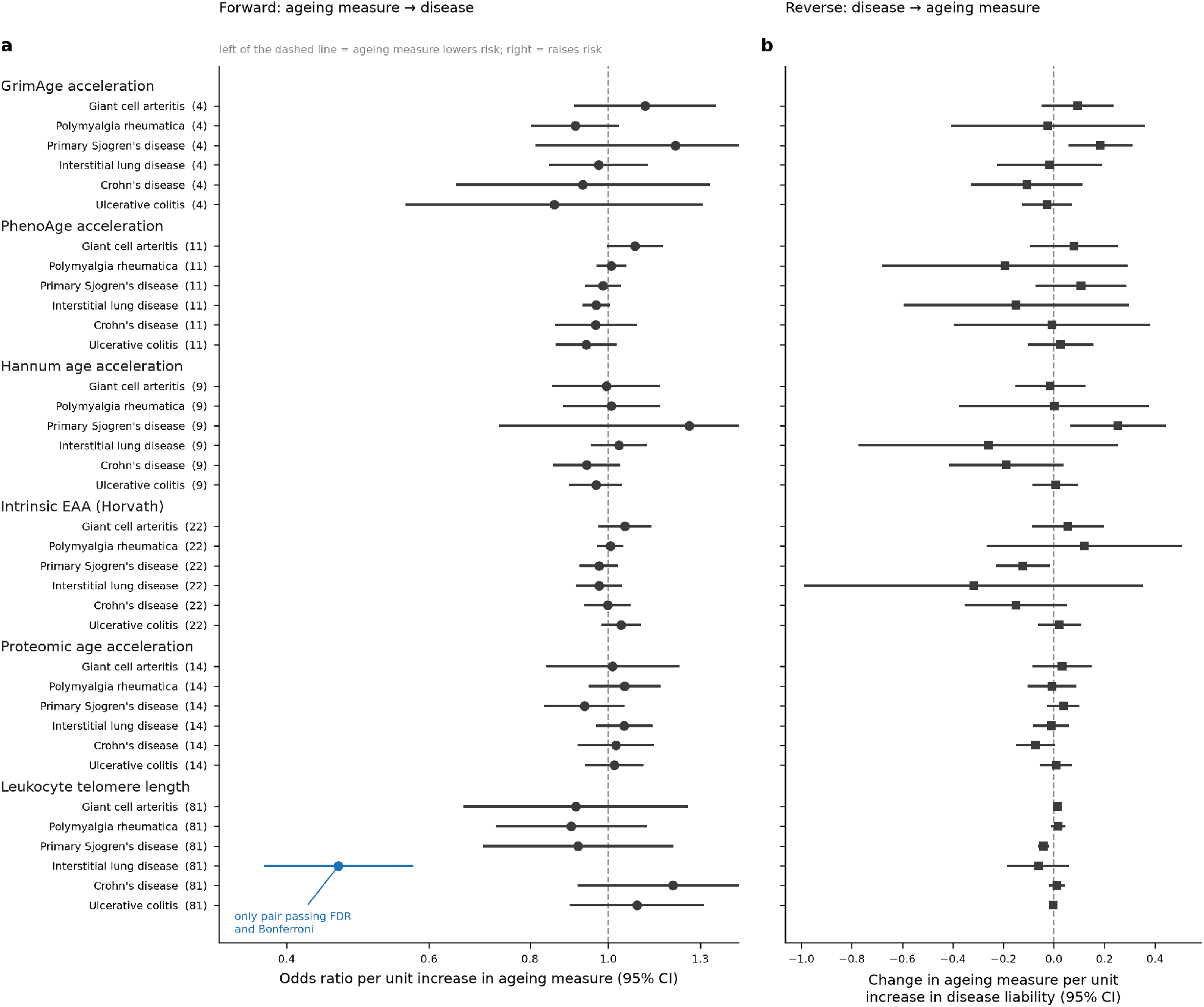
Bidirectional Mendelian randomisation across 36 exposure–outcome pairs. (a) Forward direction: inverse-variance weighted odds ratio per unit increase in each ageing measure, with 95% confidence intervals; instrument counts in parentheses. Per-standard-deviation values are in the supplementary per-SD table. The blue estimate is the only pair surviving false-discovery-rate correction. (b) Reverse direction: change in the ageing measure per unit increase in disease liability. Rows are matched between panels.

The unfiltered estimate was stronger and heterogeneous: OR 0.46 (0.37–0.57), *p* = 1.5 × 10⁻¹², *Q p* = 8.0 × 10⁻⁶, *I*² = 45.5%, with the weighted mode discordant at 0.74 (*p* = 0.44). Four instruments account for this (Figure 2): rs7705526 (*TERT*), rs12637184 (*TERC*), rs429358 (the *APOE* ε4 tag variant) and rs611646 (*ATM*). Their removal attenuates the estimate by about 25%, eliminates the heterogeneity and brings all three estimators into agreement. *TERT* and *TERC* are monogenic causes of familial pulmonary fibrosis, and *APOE* has no established role in telomere biology, so these are instruments with plausible routes to lung disease other than through circulating telomere length.

**Figure 2.**
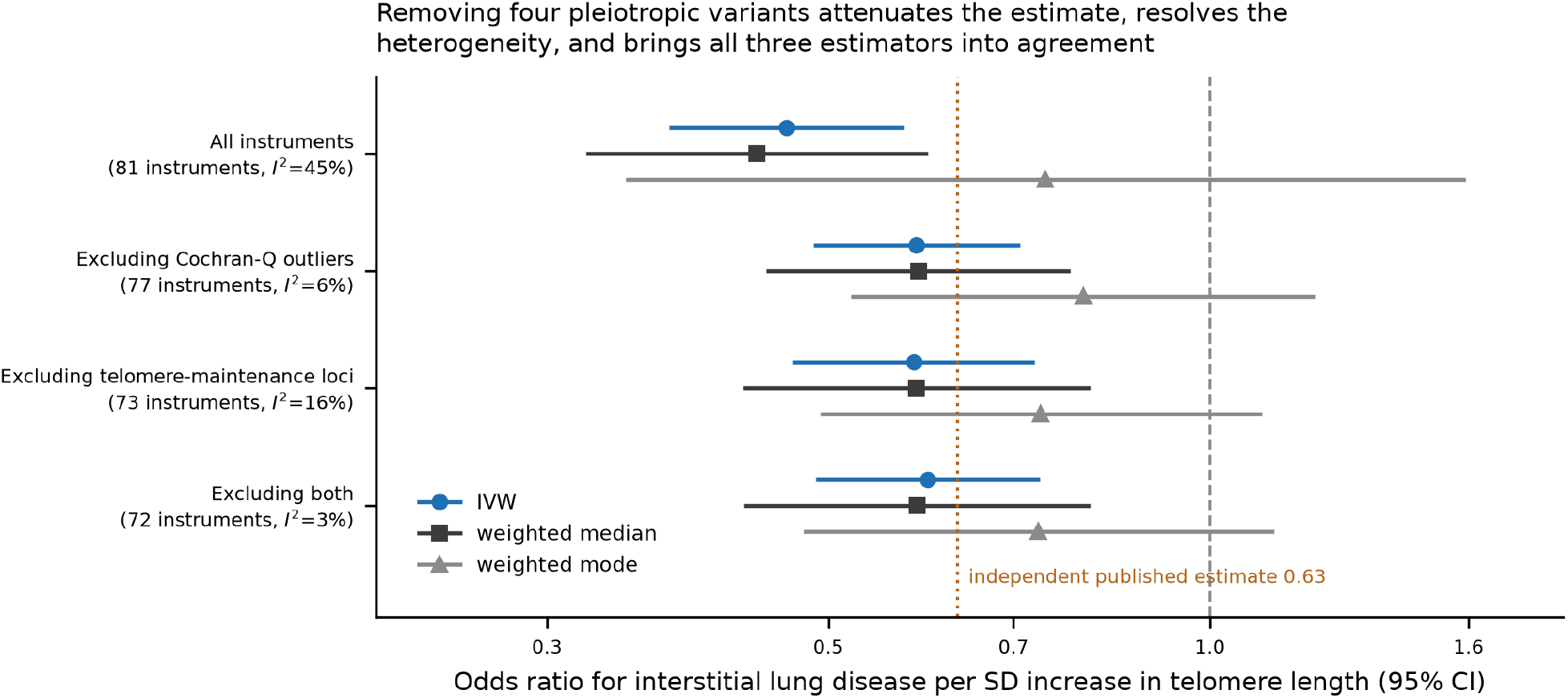
Pleiotropic instruments inflate the telomere–ILD estimate. Odds ratio for interstitial lung disease per standard deviation increase in leukocyte telomere length, under four instrument sets, by three estimators. Removing four Cochran’s *Q* outliers (*TERT*, *TERC*, *APOE*, *ATM*) attenuates the estimate by about 25%, reduces *I*² from 45% to 6%, and brings the three estimators into agreement. Excluding all eight instruments at known telomere-maintenance or pulmonary-fibrosis loci gives a similar result. The dotted line marks the independently published estimate [11].

We tested this directly by annotating all 81 instruments against telomere-maintenance and pulmonary-fibrosis loci. Eight fall within 500 kb of such a gene. Excluding the whole class gives OR 0.58 (0.47–0.73), *p* = 1.5 × 10⁻⁶ on 73 instruments (*I*² = 16.4%); excluding both the class and the *Q* outliers gives OR 0.60 (0.49–0.74) on 72 instruments (*I*² = 3.2%). The effect does not depend on those loci, but it is stronger within them (class-only OR 0.32, *Q p* = 5 × 10⁻⁸), consistent with a direct lung pathway contributing to the unfiltered pooled estimate. The attenuated estimate is unchanged by HLA exclusion and agrees closely with the independently published figure of 0.63 [11]. Seventy-three of 81 instruments were correctly oriented on the approximate directionality check. *I*²_GX was 99.4%, indicating no NOME violation and supporting the reliability of MR-Egger here; we note nonetheless that a null Egger intercept (*p* = 0.15) does not exclude balanced pleiotropy.

Because the effect allele lengthens telomeres, OR < 1 indicates that shorter telomeres — a biologically older state — increase ILD risk (Table 3).

**Table 3.** Leukocyte telomere length on interstitial lung disease, by instrument set and estimator. Odds ratios per standard deviation of telomere length.

| Instrument set | n | IVW (95% CI) | $p$ | Weighted median | Weighted mode | $I^2$ | $Q\ p$ |
| --- | --- | --- | --- | --- | --- | --- | --- |
| All instruments | 81 | 0.46 (0.37-0.57) | $1.5 \times 10^{-12}$ | 0.44 | 0.74 | 45.5% | $8.0 \times 10^{-6}$ |
| **Excluding $Q$ outliers (primary)** | **77** | **0.59 (0.49-0.71)** | **$2.7 \times 10^{-8}$** | **0.59** | **0.79** | **6.3%** | **0.32** |
| Excluding telomere-maintenance loci | 73 | 0.58 (0.47-0.73) | $1.5 \times 10^{-6}$ | 0.59 | 0.74 | 16.4% | 0.12 |
| Excluding both | 72 | 0.60 (0.49-0.74) | $8.8 \times 10^{-7}$ | 0.59 | 0.73 | 3.2% | 0.40 |
| Telomere-maintenance loci only | 8 | 0.32 (0.17-0.62) | $7.2 \times 10^{-4}$ | 0.21 | — | — | $4.9 \times 10^{-8}$ |
| All instruments, HLA excluded | 80 | 0.46 (0.37-0.57) | $1.2 \times 10^{-12}$ | — | — | — | — |

MR-Egger intercept $p = 0.15$ (all instruments); $I^2_{GX} = 99.4\%$ ; 73 of 81 instruments correctly oriented on the approximate directionality check. $Q$ outliers: rs7705526 (*TERT*), rs12637184 (*TERC*), rs429358 (*APOE*), rs611646 (*ATM*).

### Replication in independent idiopathic pulmonary fibrosis samples

The telomere–lung effect replicated in both independent samples (Figure 3, Table 4). In 5,159 clinician-diagnosed European IPF cases [23] the outlier-robust estimate was **OR 0.18 per SD (95% CI 0.12–0.25), *p* = 5.0 × 10⁻²³** on 76 instruments. In 1,210 East Asian IPF cases [24] it was OR 0.54 (0.35–0.83), *p* = 0.005 on 58 instruments, with no residual heterogeneity (*Q p* = 0.79, *I*² = 0%). Neither dataset shares participants with FinnGen, and neither uses UK Biobank as a case source, so neither is circular with respect to the exposure GWAS. Allele frequencies in the European replication sample matched the exposure sample to *r* > 0.999.

**Figure 3.**
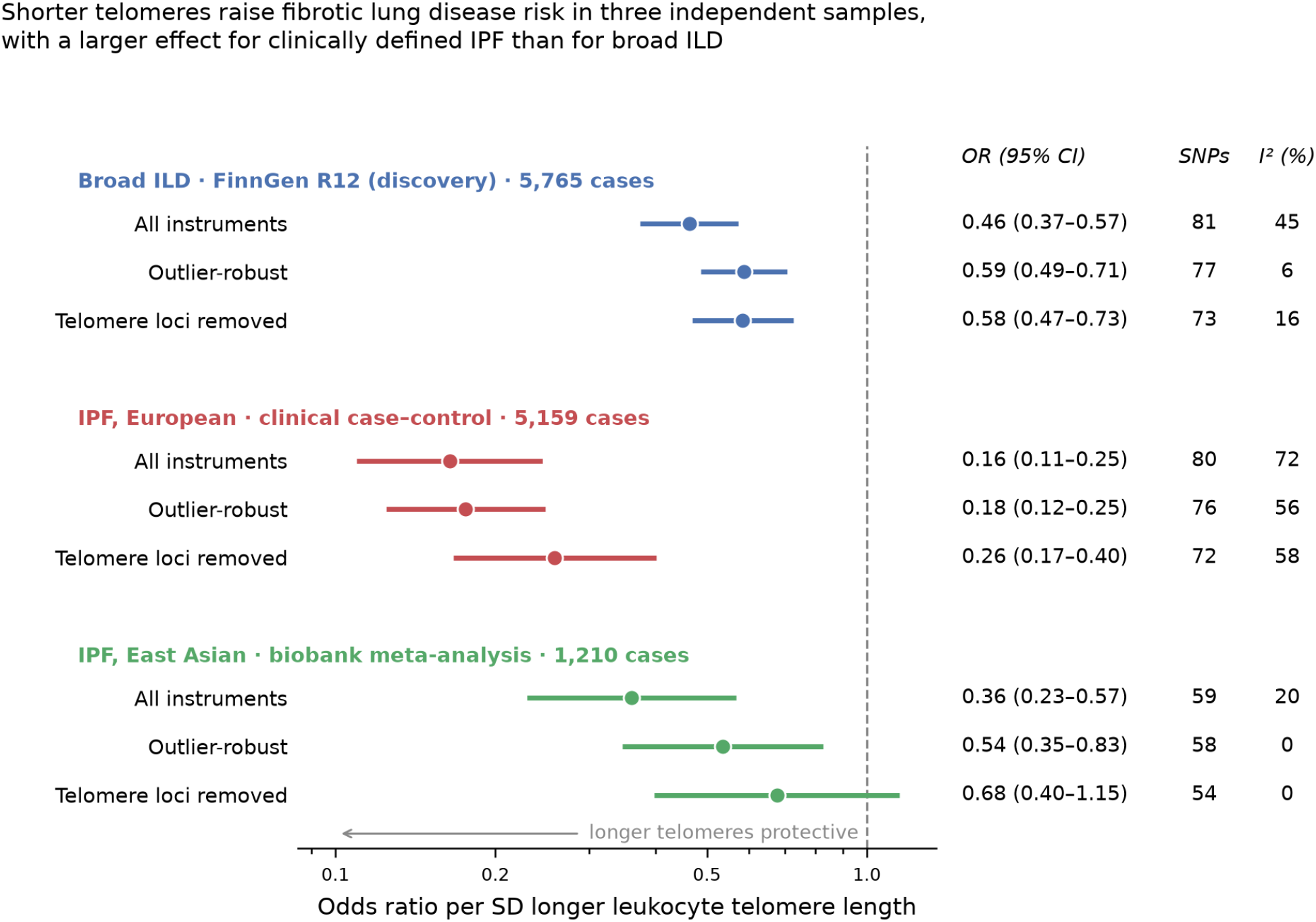
The telomere–lung effect replicates in two independent samples and is larger for a narrower case definition. Odds ratio for fibrotic lung disease per standard deviation increase in leukocyte telomere length, in the FinnGen discovery sample (broad registry ILD endpoint) and in two independent IPF case-control datasets — one of 5,159 clinician-diagnosed European cases [23], one of 1,210 East Asian cases [24] — under three instrument sets each. Neither replication sample shares participants with FinnGen or uses UK Biobank as a case source. The European IPF estimate is three times larger than the broad-ILD estimate (*z* = 6.0, *p* = 1.8 × 10⁻⁹). Removing all eight telomere-maintenance and pulmonary-fibrosis loci leaves the European IPF effect at *p* = 1.6 × 10⁻⁹. Lower odds ratios indicate stronger protection by longer telomeres.

**Table 4.**
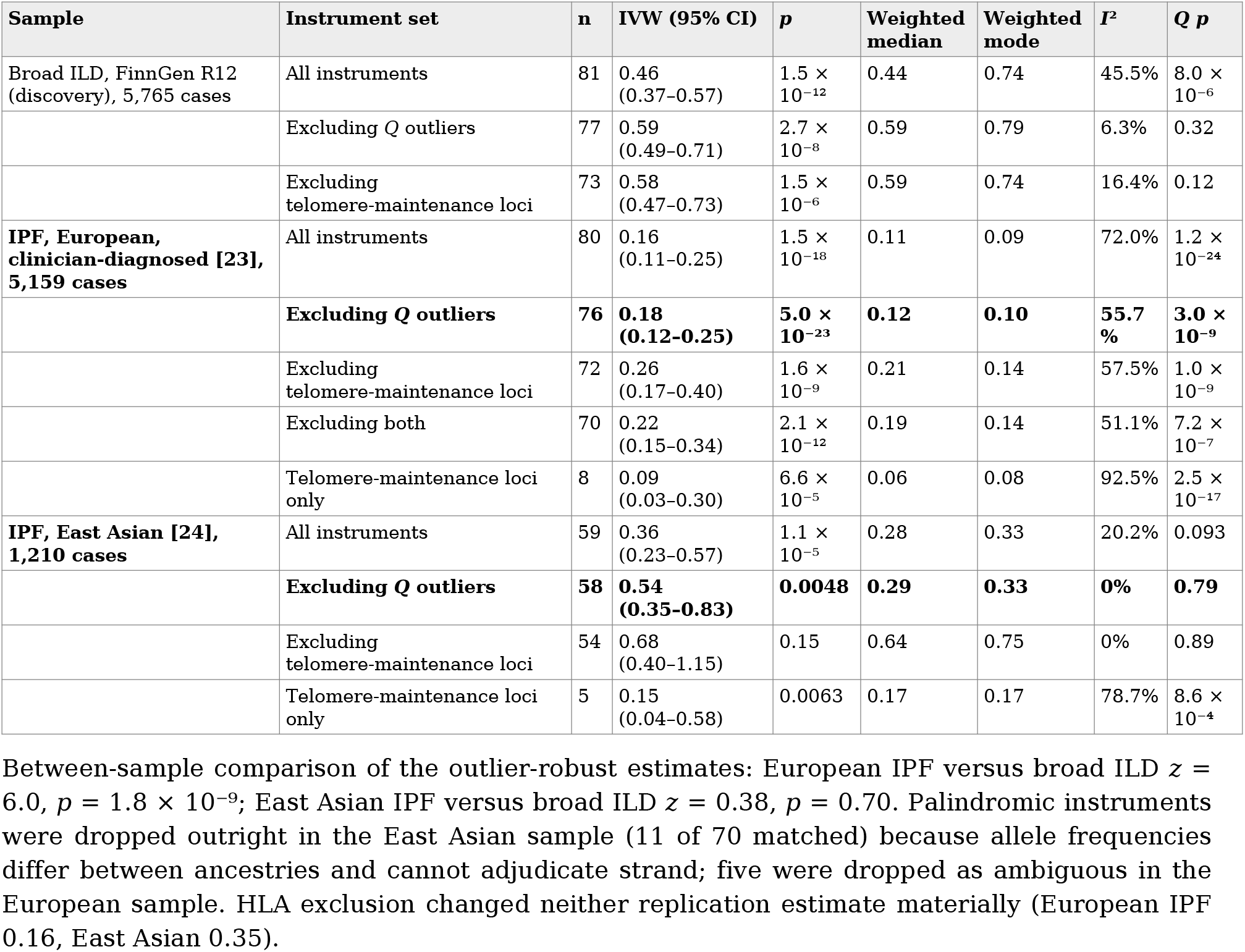
Replication in independent idiopathic pulmonary fibrosis samples. Odds ratios per standard deviation of leukocyte telomere length. Neither replication sample shares participants with FinnGen, and neither uses UK Biobank as a case source.

The European estimate of 0.18 is close to the value of 0.17 obtained by meta-analysis in an earlier telomere–IPF Mendelian randomisation [26] (reported there as OR 5.81 per SD *shorter* telomere). The agreement is reassuring but is not independent evidence: the seven case-control collections pooled in our European replication sample [23] include the collection that served as that study’s replication cohort. Our estimate differs in resting on 80 instruments rather than seven, in a larger case set, and in supporting the locus-class analysis below, which a seven-variant design cannot.

### The effect does not depend on telomere-maintenance loci

Repeating the leave-class-out cascade in the European IPF sample, exclusion of all eight instruments within 500 kb of a telomere-maintenance or pulmonary-fibrosis gene left OR 0.26 (0.17–0.40), *p* = 1.6 × 10⁻⁹ on 72 instruments; excluding the *Q* outliers as well gave OR 0.22 (0.15–0.34) on 70 instruments. This is the decisive test of the objection that the result might be carried by *TERT* and *TERC* acting on lung epithelium rather than through circulating telomere length: in a clinically phenotyped sample the effect survives removal of that entire locus class at *p* < 10⁻⁸.

This test has not previously been possible. The established telomere–IPF estimate rests on seven variants [26], six of which lie at telomere-maintenance loci (*TERC*, *TERT*, *NAF1*, *STN1*/ *OBFC1*, *ZNF208*, *RTEL1*); that instrument set cannot distinguish an effect of telomere length from an effect of telomerase-pathway genes, because the two are nearly collinear within it. Our class-only estimate of OR 0.09 (0.03–0.30) closely matches the value of 0.08 that study obtained in its replication cohort, confirming that its estimate is effectively a telomere-maintenance-locus estimate. The class-excluded estimate of 0.26, carried by 72 instruments with no known role in telomere maintenance or fibrosis, is the new quantity: it isolates the component of the effect attributable to telomere length itself. In the East Asian sample the corresponding estimate was directionally consistent but not significant (OR 0.68, 0.40–1.15, *p* = 0.15 on 54 instruments), as expected with one-fifth the case count and weaker instrument transfer.

### The causal effect is three times larger for IPF than for broad ILD

The European IPF estimate differs from the discovery broad-ILD estimate at *z* = 6.0, *p* = 1.8 × 10⁻⁹; the East Asian estimate does not (*z* = 0.38, *p* = 0.70). FinnGen’s `ILD_ENDPOINTS` aggregates hypersensitivity pneumonitis, sarcoidosis-associated and drug-induced disease together with fibrotic disease, so a mechanism specific to fibrosis is diluted by case mix. The direction and magnitude of the discrepancy are what non-differential outcome misclassification predicts, and they imply that our discovery estimate — and, by the same argument, published MR estimates built on broad registry ILD endpoints — understate the effect on fibrotic disease.

### An outlier instrument becomes a mechanistic observation

rs7705526 (*TERT*) was a Cochran-*Q* outlier in all three samples and in the same direction each time, its Wald ratio exceeding the pooled estimate by a factor of 2.6 to 4.6 (−4.56 in European IPF, −3.70 in East Asian IPF, against pooled log odds ratios of −1.74 and −0.63). An instrument whose effect on disease is consistently larger than its effect on the measured exposure predicts is the expected signature of a second, tissue-local pathway: telomerase acting on alveolar epithelial progenitors over and above its effect on leukocyte telomere length. Replication of that pattern in three samples that share no participants makes a chance explanation implausible. rs611646 (*ATM*) was likewise an outlier in both European samples, in the opposite direction, consistent with independent pleiotropy through DNA-damage response. rs429358 (*APOE*) was an outlier only in the broad ILD endpoint and not in either IPF sample, so its contribution appears specific to the heterogeneous registry phenotype.

Residual heterogeneity remained high in the European IPF sample (*I*² = 51–58% across instrument sets) and is not removable by the exclusions tested. We therefore report the weighted median (0.12–0.19 across sets) and weighted mode (0.09–0.14) alongside IVW; all three agree in direction and all exclude the null, but the spread between them bounds how precisely the magnitude can be stated. The effect on IPF is therefore large and robustly non-null, with a magnitude best expressed as a range of approximately 0.1 to 0.3 per SD rather than as a single point estimate.

### Other exposure–outcome pairs

The remaining 35 primary pairs had FDR-adjusted *p* > 0.7 (Figure 1a). The smallest uncorrected *p*-values were PhenoAge acceleration on GCA (OR 1.48 per SD, 95% CI 0.98–2.24; OR 1.08 per year, 1.00–1.17; *p* = 0.061) and PhenoAge acceleration on ILD (OR 0.83 per SD, 0.68–1.02; OR 0.97 per year, 0.93–1.00; *p* = 0.079). For the study’s motivating hypothesis, GrimAge acceleration on GCA gave OR 1.52 per SD (0.68–3.39); OR 1.11 per year (0.91–1.36), *p* = 0.31, on four instruments. Applying outlier-robust estimation across all pairs removed 23 instruments from 15 of the 36 pairs and left the number of pairs surviving correction unchanged at one.

Neither any methylation clock nor proteomic age acceleration showed an effect on ILD, the outcome on which telomere length acts most strongly; the six ageing measures did not behave as substitutes for one another for any outcome tested.

### Reverse direction

All 36 pairs were re-analysed with disease liability as the exposure, instrumented from FinnGen lead variants. Instrument counts are the limiting factor: polymyalgia rheumatica is instrumented by a single variant for four of six outcomes, forcing a bare Wald ratio; GCA by two; Sjögren’s disease by three or four; ILD by five to eight; Crohn’s disease by six or seven; ulcerative colitis by 31 or 32.

Detection limits make the consequence explicit. The smallest detectable effect ranges from 0.18 to 0.78 SD of the ageing measure per unit log-odds of disease liability, against observed effects of 0.015 to 0.115 SD. **No reverse pair could have detected its own observed effect size.** We therefore do not conclude that reverse causation is absent; we report that this design cannot test it at informative precision, with the bounds above.

One reverse pair passed correction: primary Sjögren’s disease on telomere length, β = −0.042 SD per unit log-odds liability (SE 0.012), *p* = 3.5 × 10⁻⁴, FDR *p* = 0.013 (Figure 1b). It is attributable to a single variant (below). For ILD specifically the reverse estimate was small and null, so the forward telomere-to-ILD effect is not explained by disease shortening telomeres — though that reassurance is itself bounded by the detection limit for that cell.

### Influence of a single HLA-region instrument

Four instruments lie in the extended HLA region: rs887466 (GrimAge), rs3093956 (Hannum-age), rs6914699 (IEAA) and rs2763979 (telomere length). Their influence is concentrated.

In the forward direction, GrimAge acceleration on Sjögren’s disease gave OR 1.21 (*p* = 0.35) with extreme heterogeneity (*Q* = 27.3, *p* = 5.2 × 10⁻⁶). Single-variant Wald ratios show why: rs887466 gives +0.74 (SE 0.13) against −0.12, −0.08 and +0.11 for the other three instruments. Excluding rs887466 moves the estimate to OR 0.98 (*p* = 0.83).

In the reverse direction, the Sjögren’s result above rests on rs9267092 (FinnGen Sjögren’s *p* = 2.4 × 10⁻⁹⁶). Excluding it leaves β = −0.049 (SE 0.028), *p* = 0.073 on three instruments, whose individual estimates disagree among themselves.

The telomere-to-ILD effect was unchanged by HLA exclusion. HLA pleiotropy in this design therefore does not inflate estimates indiscriminately: it specifically creates apparent associations between ageing measures and autoimmune outcomes, which is the comparison the ageing–immunity hypothesis rests on.

### Detection limits

No pair had 80% power to detect an OR of 1.20 per SD at the corrected threshold, and none at 1.10 (Figure 3). Minimum detectable ORs ranged from 1.29 (IEAA on ulcerative colitis) to 5.24 (GrimAge on GCA); the GCA analyses required 1.78–5.24. Since biological-ageing effects in this literature are of the order 1.05–1.30 per SD, the 35 null results bound the effect size rather than exclude an effect. The one detected pair had a detection limit of 1.38 against an observed effect corresponding to 1.70 in the harmful direction.

Extending the GCA analysis to adequate power is not feasible with realistic data. Holding controls at the current count, detecting an OR of 1.10 per SD at the corrected threshold would require between 56,180 GCA cases (IEAA, the best-instrumented exposure) and 3,035,766 (GrimAge); at OR 1.20 the range is 14,159 to 149,324. The largest GCA genome-wide association studies assembled to date contain a few thousand cases [13,14].

### Mediation through telomere length

Seven taxa yielded 9–17 instruments each (mean *F* 21–26, minimum 16.9). Because the telomere-length GWAS is large, this arm was well powered: mediated effects as small as OR 1.019–1.038 were detectable at 80% power (Figure 4).

**Figure 4.**
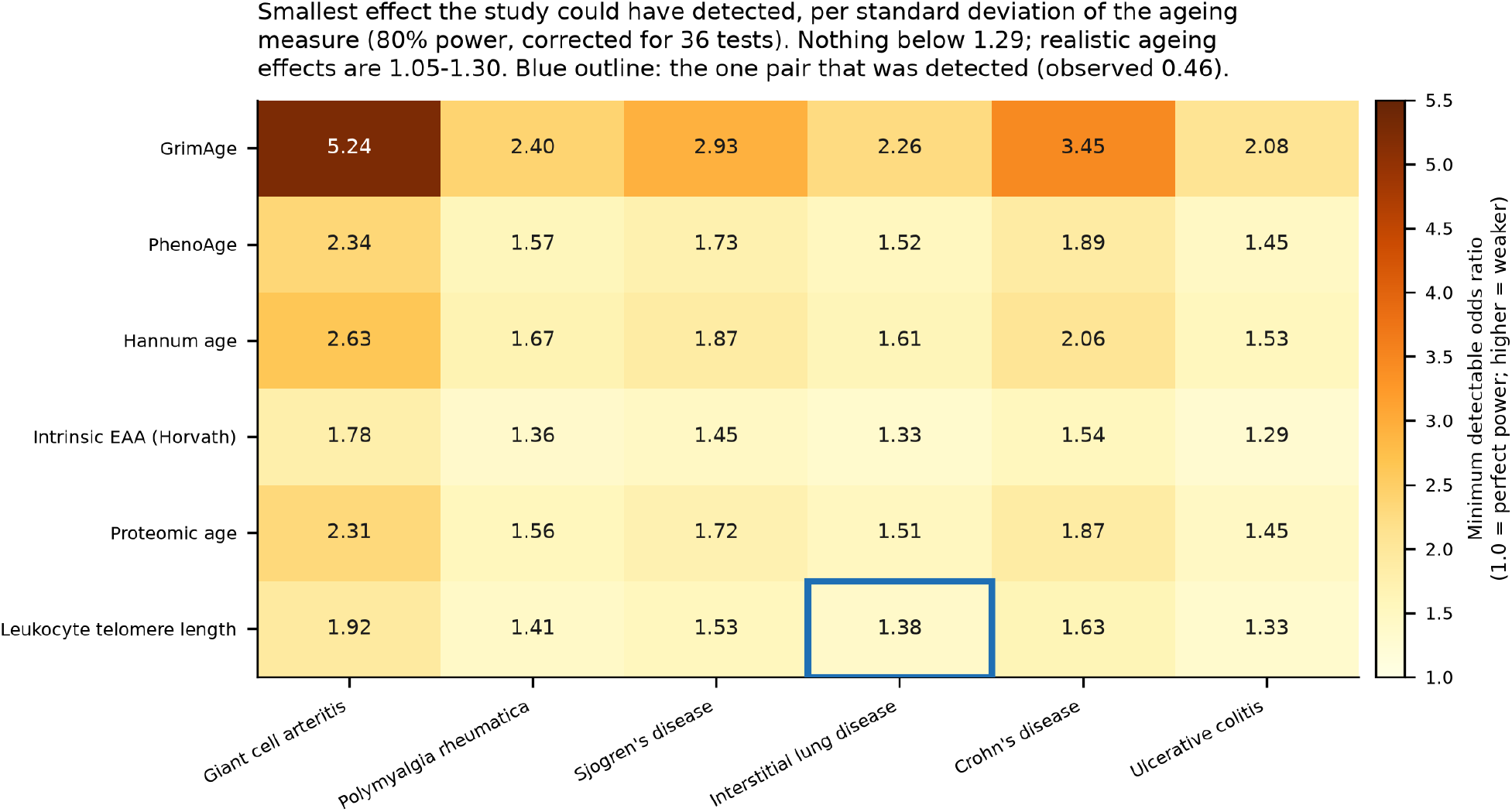
Minimum detectable effect for each pair. Smallest odds ratio per standard deviation of the ageing measure detectable at 80% power after Bonferroni correction for 36 tests. Nothing falls below 1.29, against realistic ageing effects of 1.05–1.30. Blue outline marks the one pair detected.

**Figure 5.**
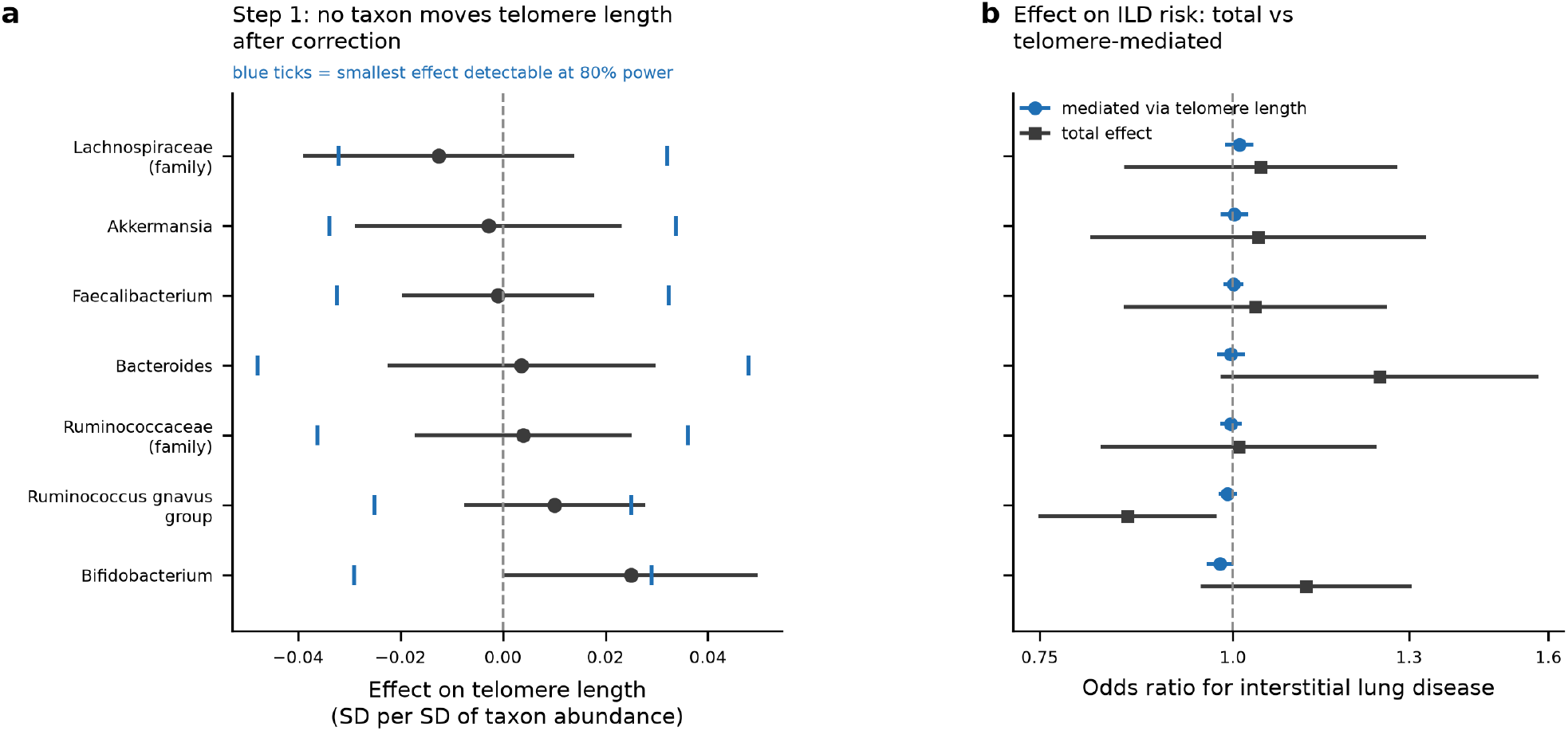
Mediation through telomere length. (a) Step 1: effect of each gut taxon on telomere length, in standard deviations per standard deviation of taxon abundance; blue ticks mark the smallest effect detectable at 80% power. (b) Total effect of each taxon on interstitial lung disease risk (grey squares) alongside the component mediated through telomere length (blue circles).

The chain failed at its first step. No taxon had a detectable effect on telomere length; every estimate fell below its own detection limit (|α| 0.0009–0.0250 SD against limits of 0.025–0.048 SD). The largest was *Bifidobacterium* (α = +0.025, uncorrected *p* = 0.047, Bonferroni *p* = 0.33). Across all 42 taxon × ageing-measure step-1 tests nothing survived FDR correction (smallest adjusted *p* = 0.76). All seven mediated effects were consequently null with narrow intervals, the widest spanning OR 0.962–1.031, excluding telomere-mediated effects greater than about 4%. One total effect was nominally significant — *Ruminococcus gnavus* group on ILD, OR 0.85, *p* = 0.021, against a Bonferroni threshold of 0.0071 — of which 4.9% was telomere-mediated and that component null; *Bacteroides* on ILD was next closest (OR 1.24, *p* = 0.070).

## Discussion

### Principal findings

Across 36 ageing–disease pairs tested in both directions, genetically proxied biological ageing was not a cause of immune-mediated disease within the effect sizes this design can detect. The single exception is mechanistically specific and now replicated: telomere attrition increases fibrotic lung disease risk, at OR 0.59 per SD of telomere length for a broad registry ILD endpoint and OR 0.18 per SD for clinically diagnosed IPF, in datasets sharing no participants with the discovery samples.

The replication addresses a central objection to any telomere–lung Mendelian randomisation, including previously published analyses: that an effect estimated from blood telomere length may instead be carried by *TERT* and *TERC* variants acting directly on lung epithelium, which would constitute pleiotropy through the measured exposure in name only. The objection applies with particular force to the conventional seven-variant instrument set [26]. Removing every instrument within 500 kb of a telomere-maintenance or fibrosis gene leaves OR 0.26 (0.17–0.40), *p* = 1.6 × 10⁻⁹ in a clinically phenotyped sample. The effect is a property of telomere length, not of a handful of fibrosis loci.

Two further results emerged from replication. The effect is three times larger for clinically defined IPF than for a broad registry ILD endpoint, which is a caution about outcome definition in biobank-based MR generally: an estimate obtained against an aggregated ILD phenotype is an attenuated estimate of the effect on fibrotic disease, and the attenuation here is not small. And the *TERT* instrument behaves as a heterogeneity outlier in the same direction in all three samples, its effect on disease exceeding what its effect on blood telomere length predicts. In a single dataset this pattern would be uninformative; reproduced in three non-overlapping samples it is evidence for a tissue-local telomerase pathway operating alongside the systemic one, and is reported here as such rather than treated only as a source of heterogeneity.

Three findings bear on how this literature should be read. First, the ageing measures are not interchangeable: telomere length acts on ILD while four methylation clocks and proteomic age act on nothing, including on ILD. Analyses that treat biological age acceleration as a single exposure are not supported by these data. Second, apparent clock–autoimmunity associations can be created by a single HLA instrument; the magnitude is not subtle, and reporting HLA-excluded estimates should be routine. Third, the headline effect itself contains a pleiotropic component that a conventional analysis would have missed: *TERT* and *TERC* instruments, whose genes cause monogenic pulmonary fibrosis through lung-epithelial telomerase insufficiency rather than through circulating telomere length, inflate the pooled estimate by about 25% and generate all of its heterogeneity. The conventional reassurance — a null MR-Egger intercept — did not detect this; outlier removal and locus annotation did.

### Comparison with the literature

The telomere–ILD relationship is not a new claim. A bidirectional MR study reported that telomere length reduces ILD risk (OR 0.63, 95% CI 0.52–0.77) with no reverse causation [11], and phenome-wide MR of telomere length has previously implicated pulmonary phenotypes [12,15]. Our broad-ILD estimate of 0.59 (0.49–0.71) agrees closely with that published figure, and the convergence is itself informative: the unfiltered estimate of 0.46 is the outlier, and the pleiotropy analysis brings our result into line with the independent one. The prior study used a registry ILD endpoint of the same kind as ours, which explains the agreement — and, in light of the case-definition result reported here, means that both estimates are attenuated with respect to fibrotic disease.

The stronger prior claim is for IPF specifically, where a seven-instrument Mendelian randomisation reported OR 5.81 per SD shorter telomere — 0.17 per SD longer — and concluded that short telomeres cause IPF [26]. Our European IPF estimate of 0.18 reproduces that figure, which is a meaningful convergence given the difference in instrument count, but is not independent of it: the case collections overlap, as stated in Methods.

Three things in the present analysis are not in that prior work. The pleiotropy dissection establishes that the effect is not carried by telomere-maintenance loci. This is the substantive advance, and it is only possible with a large instrument set: six of the seven variants used previously [26] lie at telomere-maintenance loci, so that design could not separate "short telomeres cause IPF" from "telomerase-pathway variation causes IPF" — a distinction that matters directly for whether telomere length is a tractable target or merely a marker of the pathway. Our class-only estimate reproduces the prior replication-cohort value (0.09 against 0.08), confirming that the prior estimate is effectively a locus-class estimate, while the class-excluded estimate of 0.26 on 72 unrelated instruments shows the effect persists without them. The IPF-specific replication in clinically diagnosed cases gives an effect three times larger and demonstrates that the choice of outcome phenotype, not only the choice of instrument, materially changes the answer. And the comparison class — the same outcomes tested against five further ageing measures, none of which shows an effect — reframes the finding from "ageing causes lung fibrosis" to "telomere biology specifically causes lung fibrosis". The magnitude we obtain for IPF is consistent with the large telomere effects reported for fibrotic lung disease in earlier phenome-wide and disease-specific work [12,15], though those analyses used UK Biobank outcomes and so cannot be treated as independent of the exposure GWAS.

The GCA, polymyalgia rheumatica and Sjögren’s results are, as far as we are aware, the first MR tests of biological ageing against these diseases. Their value lies in the stated bounds rather than the point estimates, and the power analysis shows that the age-dependent vasculitides cannot be settled by this design with currently available case numbers.

### Strengths

Bidirectional analysis of every pair; four estimators with differing pleiotropy assumptions; harmonisation verified by allele frequency rather than assumed; a pre-specified HLA sensitivity analysis throughout; outlier-robust re-estimation of the whole grid; locus annotation of the instruments behind the one positive finding; instruments drawn from full summary statistics rather than curated lead-variant lists; detection limits reported for every estimate in both directions and in the mediation arm, computed before the estimates were examined; and replication of the one positive finding in two datasets selected for non-overlap with both discovery samples, one of them in a different ancestry and one of them using clinician-adjudicated rather than registry-code case definitions.

### Limitations

Our European IPF replication sample is independent of the discovery samples but not of the prior literature: it subsumes the case-control collection used as the replication cohort of the earlier telomere–IPF analysis [26], so the agreement between our estimate and theirs is partly expected by construction. Only the East Asian sample is independent of all prior work, and it is the smaller and less precisely estimated of the two. Discovery outcomes are drawn exclusively from a Finnish cohort with a distinctive population history [4]; the European IPF replication sample [23] addresses this for the telomere–lung finding, but the 35 null pairs remain Finnish-only and may not generalise — non-Finnish inflammatory bowel disease summary statistics are available [6] and would allow the Crohn’s and ulcerative colitis columns to be re-tested, which we did not do. Exposure and discovery outcome samples are largely but not provably disjoint, as described in Methods; the replication samples were selected to avoid overlap with both, though we cannot exclude that a small number of control participants in the European IPF collections also appear in UK Biobank, which at a fraction well below 1% of the 472,174-participant exposure sample would bias the estimate negligibly. The cross-ancestry replication additionally assumes that variant–exposure effects transfer from European to East Asian ancestry, which is not testable here; the attenuated and non-significant estimate after removing telomere-maintenance loci in that sample (OR 0.68, *p* = 0.15) is consistent either with reduced power or with imperfect instrument transfer, and we cannot distinguish these. Residual heterogeneity in the European IPF sample remains substantial (*I*² = 51–58%) and is not removed by any exclusion tested, so the magnitude of the IPF effect is less well determined than its existence. Instruments were selected by physical distance rather than measured linkage disequilibrium, which is conservative but costs power. MR-PRESSO was unavailable; the iterative *Q* procedure implements its outlier logic but is not the same test, and no formal distortion test is reported. Directionality was assessed by an approximation to, not an implementation of, the Steiger test. GrimAge acceleration is instrumented by only four variants, leaving MR-Egger almost no power to detect pleiotropy; more severely, pleiotropy cannot be assessed by any method used for the reverse pairs with one or two instruments, and those nulls carry correspondingly less epistemic weight. The mediation analysis covers seven candidate taxa rather than a full screen, and its instruments rest on a relaxed significance threshold and a modest discovery sample [5]; its power derives from the outcome side, not the exposure side. Finally, estimator implementations were written for this study rather than taken from an established package, and although validated against an analytic standard-error relationship, they have not been benchmarked against a reference implementation.

## Conclusion

Biological ageing, as captured by four DNA-methylation clocks and a proteomic age score, shows no detectable causal effect on six immune-mediated diseases in either direction, within bounds we state explicitly for every comparison. Telomere attrition causally increases fibrotic lung disease risk — an effect that reproduces in European and East Asian case-control samples, is three times larger for clinically diagnosed IPF than for a broad registry endpoint, and, shown here for the first time, survives removal of every telomere-maintenance locus. That last result is what separates an effect of telomere length from an effect of the telomerase genes that constitute most of the conventional instrument set, and it is the main advance over the established finding [26]. It stands out as mechanistically specific rather than as one instance of a general ageing effect. Associations between epigenetic clocks and autoimmune disease in this design should be checked against HLA exclusion before interpretation, and null results in the age-dependent vasculitides should be reported with the effect sizes they can actually exclude.

## Supporting information

Strobe MR checklist

## Data Availability

All data produced in the present study are available online. This work is a secondary analysis of published genome-wide association summary statistics; no individual level data were accessed and no new data were collected. The source datasets are: FinnGen release R12 (https://www.finngen.fi/en/access_results); DNA methylation clock GWAS summary statistics, Edinburgh DataShare (https://datashare.ed.ac.uk/handle/10283/3645) and GWAS Catalog accessions GCST90014288, GCST90014289, GCST90014290, GCST90014292; leukocyte telomere length (https://figshare.com/s/caa99dc0f76d62990195); proteomic age acceleration (https://doi.org/10.17863/CAM.116389); idiopathic pulmonary fibrosis replication datasets, GWAS Catalog accessions GCST90984755 and GCST90399719; and gut microbiome GWAS summary statistics, GWAS Catalog accessions GCST90016940 till GCST90017065. All instrument tables, harmonised per-variant datasets, complete estimator output including analyses not presented in the manuscript, figures, and the full analysis code are deposited at Zenodo under the MIT licence: https://doi.org/10.5281/zenodo.23190662.

https://www.finngen.fi/en/access_results

https://datashare.ed.ac.uk/handle/10283/3645

https://www.ebi.ac.uk/gwas/publications/34187551

https://figshare.com/s/caa99dc0f76d62990195

https://doi.org/10.17863/CAM.116389

https://www.ebi.ac.uk/gwas/studies/GCST90984755

https://www.ebi.ac.uk/gwas/studies/GCST90399719

https://www.ebi.ac.uk/gwas/publications/33462485

## Declarations

### Ethics approval and consent

This study used only publicly available, de-identified summary-level data from previously published genome-wide association studies. No individual-level data were accessed and no new data were collected. Ethical approval and participant informed consent were obtained by each contributing study as described in the corresponding primary publications, including FinnGen (Coordinating Ethics Committee of the Hospital District of Helsinki and Uusimaa; consent for biobank research under the Finnish Biobank Act) and UK Biobank (North West Multi-centre Research Ethics Committee). No further ethical approval was required for the present analysis.

### Data availability

All data analysed are publicly available summary statistics; accessions are given in Table 1. Disease outcomes are from the FinnGen R12 public release. All instrument tables, harmonised per-variant datasets, estimator output and figures generated for this study are deposited at Zenodo, https://doi.org/10.5281/zenodo.23190662.

### Code availability

All analysis code is deposited at Zenodo, https://doi.org/10.5281/zenodo.23190662, under the MIT licence, including the estimator library and the scripts used to extract instrument variants from remote summary-statistic files.

### Author contributions

Sten Hanke is the sole author and is responsible for all aspects of the work: conceptualisation, methodology, software, formal analysis, data curation, investigation, visualisation, writing of the original draft, and review and editing.

## Acknowledgements

We thank the participants and investigators of FinnGen, UK Biobank, the epigenetic-clock, proteomic-age and gut-microbiome consortia, and the idiopathic pulmonary fibrosis case-control collections whose summary statistics enabled the replication analyses; and the NHGRI-EBI GWAS Catalog for curating and distributing the summary statistics used throughout.

## Reporting guideline

STROBE-MR [21]; the completed 20-item checklist is supplied as a supplementary file.

## Analytical tooling

Analysis code was written with the assistance of an AI coding tool and reviewed by the authors. All statistical results reported here were generated by the deposited code, which is available for independent verification. The authors take full responsibility for the content, the analytical choices and the interpretation.

## Supplementary files

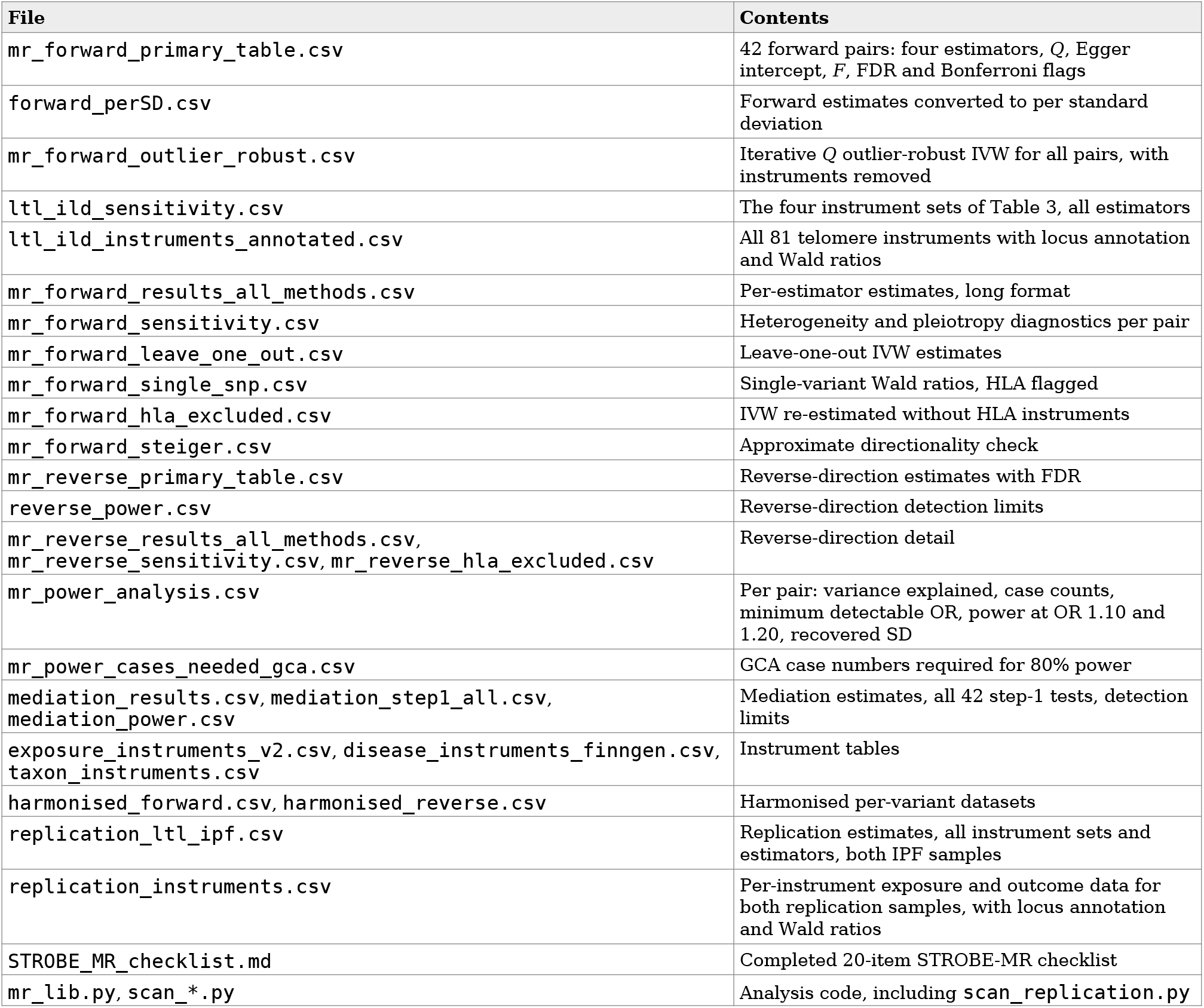

