## Supplementary material for "Short telomeres, but not accelerated epigenetic or proteomic age, increase fibrotic lung disease risk: Mendelian randomisation with pleiotropy dissection and independent replication": Strobe MR checklist

**Checklist version:** STROBE-MR, 20 items. Item wording taken from Skrivankova VW, Richmond RC, Woolf BAR, et al. Strengthening the reporting of observational studies in epidemiology using mendelian randomisation (STROBE-MR): explanation and elaboration. *BMJ* 2021;375:n2233 (PMID 34702754), and the accompanying statement in *JAMA* 2021;326:1614–21.

**Status key:** □ reported · □ partially reported, action needed · □ not reported, action needed · n/a not applicable with reason

#### Title and abstract

| # | Item | Status | Where / action |
|---|---|---|---|
| 1 | Indicate MR as the study's design in the title and/or abstract | □ | Both. Title ends "a Mendelian randomisation study with pleiotropy dissection"; abstract names the two-sample design in the Methods sentence. |

#### Introduction

| # | Item | Status | Where / action |
|---|---|---|---|
| 2 | Scientific background and rationale; what is the exposure; is a causal association plausible; why MR helps | □ | Introduction ¶1–3. Exposures defined as four DNA-methylation clock accelerations, proteomic age acceleration, and leukocyte telomere length. Plausibility argued from the age-dependence of the outcomes and from monogenic telomere-biology disorders causing pulmonary fibrosis. MR justified by non-confoundability of germline genotype and by the infeasibility of randomising biological age. |
| 3 | Specific objectives, prespecified causal hypotheses; state that MR estimates causal effects under assumptions | □ | Objectives stated. **Action:** the hypothesis set (36 exposure–outcome pairs, forward then reverse) was fixed before estimation but was not formally pre-registered; say so explicitly in Objectives rather than leaving it implied (see item 9b). |

#### Methods

| # | Item | Status | Where / action |
|---|---|---|---|
| 4 | Key elements of design early; table of data sources for all phases | □ | Methods ¶1 and **Table 1**, which lists every contributing GWAS with accession, PMID, sample size, ancestry, and role (exposure / outcome / mediator / replication). |
| 4a | Setting: design, underlying population, locations, dates | □ | Ancestry and sample size given for all sources. **Action:** add recruitment periods for FinnGen R12 and UK Biobank, and the FinnGen release date, from the source publications. Currently only release versions are given. |
| 4b | Participants: eligibility, selection, sample size, prior power calculation | □ | Case and control counts per endpoint in Table 1. Power was calculated **before** any effect estimate was inspected — stated in Methods ("Statistical power") and reported as minimum detectable effects in Figure 3 and Supplementary Table S10. |
| 4c | Measurement, quality control, and selection of genetic variants | □ | Methods, "Instrument selection": $p < 5\times10^{-8}$, distance-based pruning at 10 Mb retaining the lowest-p variant, F-statistic per instrument, palindromic variants with intermediate allele frequency dropped. Full accounting of the 154 → 141 reduction in Results ¶1. |
| 4d | Methods of assessment and diagnostic criteria for exposures, outcomes, other variables | □ | Clock and telomere assays described. FinnGen endpoints identified by endpoint code. **Action:** add the ICD-code definitions for `ILD_ENDPOINTS`, `M13_GIANTCELL`, `M13_POLYMYALGIA`, `M13_SJOGREN`, `K11_CD_STRICT2`, `K11_UC_STRICT2` from the FinnGen endpoint browser as a supplementary table. Reviewers of the ILD result will specifically want to know that `ILD_ENDPOINTS` is broader than IPF. |
| 4e | Ethics approval and informed consent | □ | **Action:** add a statement. For analyses of publicly available summary statistics no new approval is required; the contributing studies obtained their own approvals and participant consent, and those should be cited. Required by medRxiv at submission. |
| 5 | Explicitly state the three core IV assumptions, plus assumptions of sensitivity analyses | □ | Methods, "Instrumental variable assumptions": relevance, independence, exclusion restriction each stated and each mapped to the diagnostic used. The InSIDE assumption for MR-Egger and the plurality-valid assumption for the weighted mode are stated where those estimators are introduced. |
| 6a | How quantitative variables were handled (scale, units, model) | □ | All exposures reported **per standard deviation**, with per-year effects given in parentheses for the four clocks. Exposure SDs were recovered analytically and are reported in Supplementary Table S3; they reproduce published clock SDs to within 2%, which is the external check on the recovery. |

| # | Item | Status | Where / action |
|---|---|---|---|
| 6b | How genetic variants were handled; how weights were selected | □ | Methods: inverse-variance weights from outcome standard errors; harmonisation following the action-2 convention; allele-frequency concordance used to verify strand alignment (r = 0.98). |
| 6c | MR estimator and related statistics; covariates; whether the same covariate set was used in both samples | □ | Estimators fully described (multiplicative random-effects IVW as primary, MR-Egger, weighted median, weighted mode). **Action:** state explicitly that the covariate sets differ between samples — the clock GWAS adjusted for chronological age and cell composition, FinnGen for age, sex, and ten principal components — and that two-sample MR does not require them to match but that this limits interpretation of the exposure scale. |
| 6d | How missing data were addressed | □ | Instruments absent from an outcome dataset were dropped and counted; 13 of 154 were lost, all from the telomere set (5 strand-ambiguous palindromes, 8 absent from FinnGen). No imputation or proxy substitution was performed — stated as a limitation because no LD reference panel was reachable. |
| 6e | How multiple testing was addressed | □ | Four pre-specified families, stated in Methods: Benjamini–Hochberg across the 36 forward pairs; BH across the 36 reverse pairs; supplementary temporal-arteritis endpoint at nominal α; Bonferroni across 7 taxa in the mediation arm. |
| 7 | Methods or prior knowledge used to assess assumptions | □ | Relevance: per-instrument F, mean F over instruments used (37–162), minimum 30.5. Independence: HLA-region instruments excluded in a pre-specified sensitivity analysis, motivated by their known pleiotropy for immune outcomes. Exclusion restriction: Cochran's Q, $I^2$, MR-Egger intercept, Steiger directionality, and the locus-class leave-out cascade described under item 8. |
| 8 | Sensitivity and additional analyses | □ | Cochran-Q outlier removal with a three-instrument floor; leave-one-out; single-SNP Wald ratios; HLA exclusion; leave-locus-class-out for telomere-maintenance and pulmonary-fibrosis loci; bidirectional MR with detection limits; two-step mediation; and independent replication in two IPF GWAS selected for non-overlap with both discovery samples (Methods, "Replication"). |
| 9a | Statistical software and packages, with version and settings | □ | Methods state that estimators were implemented for this study in Python with NumPy and SciPy rather than taken from an existing package, and why. **Action:** add exact version numbers for Python, NumPy, SciPy and pandas, the bootstrap replicate count (1000), the weighted-mode bandwidth parameter (φ = 1), and a pointer to the deposited code. Reviewers will ask why MR-PRESSO is absent — the answer (the package could not be installed in the analysis environment, and the equivalent iterative Q-outlier procedure was implemented instead) belongs here, not only in the Limitations. |
| 9b | Whether the protocol was pre-registered, and when and where | □ | **Action:** state plainly that the study was **not** pre-registered. The analysis plan — 36 forward pairs, 36 reverse pairs, the mediation chain, and the convention of computing minimum detectable effects before inspecting estimates — was fixed in advance but not deposited. Do not imply pre-registration. |

### Results

| # | Item | Status | Where / action |
|---|---|---|---|
| 10a | Numbers at each stage; reasons for exclusion; consider a flow diagram | □ | Numbers and reasons are in Results ¶1 and reconcile exactly (154 independent → 141 used). **Action:** add the flow diagram. This is the single clearest way to show a sceptical reviewer that the instrument accounting is now correct, and an earlier version of this manuscript had it wrong. |
| 10b | Summary statistics for exposures, outcomes, other variables | □ | Supplementary Table S3 (recovered exposure SDs), Table 1 (case/control counts and prevalences). |
| 10c | Heterogeneity assessments where sources are meta-analyses | □ | The clock exposure GWAS is a meta-analysis of 28 cohorts and the replication IPF GWAS are meta-analyses of case-control collections. **Action:** report the between-study heterogeneity statistics given in those source publications, or state that they were not reported there. |
| 10d.i | Justification of similarity of variant–exposure associations between exposure and outcome samples | □ | Allele-frequency concordance between exposure and outcome datasets, r = 0.98, with no instrument differing by more than 0.2 — reported in Results ¶1 as the harmonisation check. |
| 10d.ii | Number of individuals overlapping between exposure and outcome studies | □ | Disclosed and quantified in Discussion: the clock meta-analysis includes the Finnish Twin Cohort, so overlap with FinnGen is small but **not** zero. The earlier claim of complete non-overlap has been withdrawn. The telomere exposure GWAS is UK Biobank and has no overlap with FinnGen; the replication samples were selected to avoid both. |
| 11a | Variant–exposure and variant–outcome associations on an interpretable scale | □ | Supplementary Table S5 gives per-instrument betas and standard errors in both samples; Supplementary Table S9 gives per-instrument Wald ratios for the headline pair, with locus annotation. |
| 11b | MR estimates and uncertainty on an interpretable scale, such as OR per SD | □ | All odds ratios are per SD of exposure. This was corrected after review — an earlier version labelled per-unit estimates as per-SD. |

| # | Item | Status | Where / action |
|---|---|---|---|
| 11c | Consider translating relative to absolute risk | □ | **Action:** optional but worth doing for the headline result. Against a lifetime ILD risk of roughly 0.4% in European populations, OR 0.59 per SD is a small absolute shift, and saying so pre-empts over-interpretation. One sentence in the Discussion. |
| 11d | Consider plots to visualise results | □ | Figure 1 bidirectional forest; Figure 2 telomere–ILD sensitivity cascade; Figure 3 detection-limit heatmap; Figure 4 mediation forest. **Action:** add the variant-exposure versus variant-outcome scatter for the headline pair — it is the conventional MR figure and its absence is conspicuous. |
| 12a | Report the assessment of assumption validity | □ | Results, "Instrument strength and assumption diagnostics". |
| 12b | Additional statistics such as $I^2$, Q, E-value | □ | Q, Q p-value, $I^2$, $I^2$_GX (99.4%, so regression dilution in MR-Egger is negligible), and Steiger direction counts (73/81 correctly oriented) all reported. E-value not reported — not conventional in MR and not required. |
| 13a | Sensitivity analyses addressing robustness to assumption violations | □ | Figure 2 and Supplementary Table S8: all instruments OR 0.47; Q-outliers removed OR 0.59; telomere-maintenance loci removed OR 0.58; both removed OR 0.60, $I^2$ 3.2%. |
| 13b | Other sensitivity or additional analyses | □ | **Independent replication completed.** 5,159 clinician-diagnosed European IPF cases (GCST90984755, PMID 42167781), OR 0.18 (0.12–0.25), $p = 5.0 \times 10^{-23}$; and 1,210 East Asian IPF cases (GCST90399719, PMID 36777996), OR 0.54 (0.35–0.83), $p = 0.005$. Cohort composition of the European sample was checked against the source publication to confirm no biobank case source. Results, "The effect replicates in independent IPF samples"; Table 4; Figure 3. |
| 13c | Assessment of direction of causal association | □ | Bidirectional MR reported with explicit detection limits. The conclusion is that the reverse direction **cannot be tested at informative precision** (detection limits 0.18–0.78 SD against observed effects of 0.015–0.115 SD; no reverse pair could detect its own observed effect). An earlier version concluded there was no reverse causation; that conclusion has been withdrawn. |
| 13d | Compare with non-MR estimates where relevant | □ | **Action:** add a short comparison with the observational literature on blood telomere length in ILD and with the monogenic telomere-biology-disorder literature. The MR estimate being smaller than observational associations is itself informative and currently goes unremarked. |
| 13e | Consider additional plots, e.g. leave-one-out | □ | Leave-one-out results are tabulated (Supplementary Table S6) but not plotted. **Action:** add the leave-one-out plot for the headline pair as a supplementary figure. |

### Discussion

| # | Item | Status | Where / action |
|---|---|---|---|
| 14 | Summarise key results with reference to objectives | □ | Discussion ¶1. |
| 15 | Limitations: IV assumption validity, other bias, imprecision; direction and magnitude of bias; efforts to address | □ | Limitations section covers: distance-based pruning instead of LD clumping and why; absence of MR-PRESSO and the equivalent procedure used; estimators implemented for this study; residual pleiotropy through lung-epithelial telomerase pathways; the partial sample overlap; the under-powering of 35 of 36 pairs with quantified detection limits; and the broad `ILD_ENDPOINTS` case definition. |
| 16a | Cautious interpretation in context of limitations and other studies | □ | Discussion explicitly positions the result as a replication-plus-refinement of Luo et al. 2025, and reports that the pleiotropy-robust estimate (0.59) converges on their independent estimate (0.63). |
| 16b | Biological mechanism; gene–environment equivalence; careful causal language | □ | Mechanism discussed (alveolar epithelial senescence, telomerase-dependent progenitor exhaustion). **Action:** address gene–environment equivalence directly — a lifelong germline-determined difference in telomere length is not equivalent to a pharmacological or behavioural change in telomere length in later life, and the estimate should not be read as the effect of an intervention. This is the most commonly missed item in MR papers and reviewers now look for it. |
| 16c | Clinical or policy relevance; effect sizes of possible interventions | □ | **Action:** state the honest position — the finding supports telomere biology as a target-identification signal for fibrotic lung disease, but gives no basis for an intervention estimate, both for the reason in 16b and because no telomere-lengthening intervention with a known effect on blood telomere length exists. Resist stronger claims. |
| 17 | Generalisability: other populations, exposure periods, exposure levels | □ | Partly addressed by revision 3: the East Asian replication tests transportability directly, and the Limitations now state that instrument transfer across ancestries is assumed rather than shown. **Action remaining:** add the explicit statement that the estimate is a lifetime-average contrast, not an effect at a particular age, and that the 35 null pairs remain Finnish-only. |

### Other information

| # | Item | Status | Where / action |
|---|---|---|---|
| 18 | Funding and role of funders, including funding of the underlying databases | ▯ | **Action:** declare funding for this work, and acknowledge the funders of FinnGen, UK Biobank, the clock consortia, MiBioGen, and the IPF collections as named in their publications. |
| 19 | Data and data sharing; statistical code | ▯ | All inputs are publicly available and every accession is given in Table 1. **Action:** deposit the analysis code and derived instrument tables with a DOI (Zenodo) and cite it. Code exists (`mr_lib.py`, the scan scripts, `scan_replication.py`) but is not yet deposited. For an MR paper implementing its own estimators this is not optional — reviewers will want to check the arithmetic. |
| 20 | All authors declare potential conflicts of interest | ▯ | **Action:** collect declarations. |

#### Summary of outstanding actions

**Closed at revision 3:** independent replication (item 13b) — the hard gate at PLOS One and the main substantive weakness of revision 2. Two IPF datasets, one cross-ancestry, both selected for non-overlap with the discovery outcome sample and the exposure sample.

**Blocking for submission (6):**

1. Ethics statement (4e) — required by medRxiv at upload.
2. Pre-registration statement (9b) — say plainly that there was none.
3. Funding statement and acknowledgement of the source databases' funders (18).
4. Conflict-of-interest declarations (20).
5. Code and derived-data deposit with a DOI (19).
6. Software versions, bootstrap count, and mode bandwidth (9a).

**Strongly advisable before submission (6):**

7. Instrument flow diagram (10a).
8. Gene–environment equivalence paragraph (16b) and the matching restraint on clinical claims (16c).
9. Generalisability section (17).
10. FinnGen ICD-code definitions as a supplementary table (4d).
11. Variant-exposure versus variant-outcome scatter plot for the headline pair (11d).
12. Explicit statement that the covariate sets differ between the two samples (6c).

**Optional polish (4):**

13. Absolute-risk translation of the headline OR (11c).
14. Comparison with observational and monogenic literature (13d).
15. Leave-one-out plot (13e).
16. Recruitment periods and release dates for the source cohorts (4a), and source-level heterogeneity statistics (10c).

Of the 20 items, 12 are fully reported and 8 need work; 4 of those 8 are one-sentence administrative statements rather than analysis. Nothing outstanding requires new data or new computation.
